# Regional differences in vaccine uptake and serological responses to vaccine and circulating strains of H1N1 viruses among patients with confirmed influenza

**DOI:** 10.1101/2020.10.03.20203042

**Authors:** Ashley L. Fink, Hsuan Liu, Kathryn Shaw-Saliba, Thomas Mehoke, Jared Evans, Zhen-Ying Liu, Mitra Lewis, Lauren Sauer, Peter Thielen, Kuan-Fu Chen, Richard Rothman, Sabra L. Klein, Andrew Pekosz

## Abstract

**Background:** Seasonal epidemics of influenza are often characterized through national or international surveillance efforts to determine vaccine efficacy and vaccine strain selection, but they do not provide detailed information about local variations in factors that can influence influenza cases and disease severity.

**Methods:** Surveillance for influenza like illness was performed in Emergency Medicine Departments in Taipei, Taiwan and Baltimore, Maryland during the winter of 2015-16. Detailed demographic and clinical data were obtained. Nasal swabs or washes were collected for influenza virus diagnosis, sequencing and isolation. Serum was collected to determine neutralizing antibody levels.

**Results:** H1N1 viruses dominated both sites, but more influenza cases occurred in Taipei compared to Baltimore. H1 HA clade diversity was greater in Taipei. Vaccination rates were lower in Taipei than Baltimore, but vaccination was associated with an increase in serum neutralizing antibodies to recent H1N1 strains in Taipei, but not Baltimore. There was a higher level of preexisting immunity to circulating H1N1 strains in Baltimore.

**Conclusions:** Regional differences in preexisting immunity and H1N1 strain circulation may have contributed to the vastly different 2015-16 influenza seasons in Taipei and Baltimore and suggest immune responses to vaccination can be affected by the degree of preexisting immunity in the population.

---

Influenza causes disease in the human population and the specific strain of virus that is circulating, the levels of preexisting immunity to circulating viruses, vaccine uptake, vaccine strain match to circulating virus strains, and the presence of mutations that modulate virus fitness all contribute to the magnitude of seasonal cases and disease severity [1, 2]. Other factors impact the susceptibility to influenza in a strain independent manner, including age, immune status, pregnancy, gender, and the presence of co-morbidities [2-4].

The 2015-16 influenza season in the Northern hemisphere was dominated by H1N1 Influenza A virus, with significant differences in the number of cases and disease severity across different countries [5]. In contrast, East and Southeast Asia experienced a particularly strong influenza season [5]. Serological assays using ferret antiserum indicated circulating H1N1 viruses were antigenically similar to the 2015-16 vaccine strain [6-9]. Surveillance for influenza-like illness (ILI) in 2015-16 was performed in the Emergency Departments (ED) of Chang Gung Memorial Hospital (CGMH), Taipei, Taiwan and the Johns Hopkins Medical Institution (JHMI), Baltimore, Maryland. The influenza season in Taipei was more severe than in Baltimore and factors including differences in circulating virus strains, vaccination rates, serum neutralizing antibody levels after vaccination, antibody cross reactivity between vaccine and circulating strains, and levels of preexisting immunity contributed to the different influenza seasons at the two surveillance sites.

## Materials and Methods

### Human sample collection and ethics

The human subjects protocol was reviewed and approved by the JHMI Institutional Review Board (IRB) and the CGMH IRB (IRB00052743). This protocol allowed for the enrollment of influenza positive, negative, and asymptomatic patients from the JHMI and CGMH ED with informed consent. For each patient, nasal pharyngeal (NP) swab or nasal wash (NW) and 10 ml of blood were collected at enrollment (i.e., time of presentation and positive influenza test, ≤ 7 days after the onset of symptoms). NP swabs were tested for influenza A virus using the CepheidGeneExpert© test. Blood was collected in Vacutainer tubes (BD), serum was separated and stored at −70°C. The patient’s basic demographics (i.e., age, ethnicity, race, gender, co-morbidities, and medications) were collected using data collection forms. Disease severity data (i.e., use of oxygen, hospital admission, disease outcomes) were collected during a follow up conversation and by chart review. For vaccination status, patients were asked if they received an influenza vaccine for the current season (2015-2016).

### Virus sequencing and phylogenetic analysis

Viral RNA isolation and sequencing was performed as previously described [10, 11] and the phylogenetic analysis was performed as previously described [12] by comparing the full-length nucleotide sequence of the hemagglutinin (HA) open reading frame from representative strains of all clades of H1N1 2009 pandemic viruses retrieved from the Global initiative on sharing all influenza data (GISAID). Trees were viewed and colored using FigTree and HA clades were identified by adding known clade references to these trees and identifying branches for the 6B, 6B.1, and 6B.2 clades. Clade 6B.1 was defined by HA1 amino acid substitutions S84N, S162N and I216T; and clade 6B.2 was defined by HA1 amino acid substitutions V152T and V173I and HA2 amino acid substitutions E164G and D174E. Sequences generated in the study were submitted to GenBank NCBI Influenza Virus Database (**Supplementary Table 1)**.

### Cell cultures

Madin-Darby canine kidney (MDCK) cells were used to grow all viruses and were cultured in Dulbecco’s modified Eagle medium (DMEM; Sigma) with 10% fetal bovine serum (Gibco), 100 μg/ml of penicillin and 100 μg/ml of streptomycin (Quality Biological), and Glutamax (Gibco) at 37°C and 5% CO_2_. Human nasal epithelial cell (hNEC) cultures were isolated from non-diseased tissue after endoscopic sinus surgery for non-infection related conditions and grown at air-liquid interface (ALI). The hNEC isolation, differentiation and culture conditions have been described in detail previously 125 [13].

### Vaccine viruses

A recombinant influenza virus encoding the HA protein derived from the sequence of A/Michigan/45/2015-like egg-adapted virus (A/Mich/45/15; GISAID Accession no.: EPI685579) was used as the A/Mich/45/15 vaccine virus for serology assays. The HA protein has an egg-adapted mutation at position 223 (glutamine (Q) to arginine (R); Q223R) on the HA protein. A/Mich/45/15 virus was generated using a 12-plasmid influenza reverse genetic system [14, 15] with plasmids encoding the HA and the neuraminidase (NA) of the A/Mich/45/15-like virus and the remaining 6 viral gene segments derived from A/California/7/2009 (A/Cal/7/09) as described previously [16].

### Viral isolates from clinical samples used for serology and growth analysis

Fluids from NP swabs or NW from influenza A virus positive individuals were used to isolate viruses by inoculating primary, differentiated hNEC cultures as previously described [12]. A representative viral isolate from each clade was used for serology assays and growth analysis on hNECs. The representative viral isolates used, include: 142 A/Baltimore/0026/2016 (JHMI 6B.1), A/Linkou/0032/2015 (CGMH 6B.1), 143 A/Linkou/0029/2015 (CGMH 6B.2), and A/Linkou/0030/2015 (CGMH 6B).

### Virus growth curves and TCID_50_ assay

Low-multiplicity of infection (MOI) growth curves were performed in quadruplicate by infecting hNECs in 24-well Transwell© inserts at an approximate MOI of 0.1 50% tissue culture infectious units (TCID_50_) per cell as described previously [15, 16].

### Ferret antisera

Ferrets (male, Triple F Farms, Sayre, PA) were lightly anesthetized with isoflurane and intranasally inoculated with 10^6^ TCID_50_ in 1 ml PBS of A/Baltimore/0026/2016 (6B.1) (two ferrets) or A/Linkou/0029/2015 (6B.2) (two ferrets). After 21 days, animals were sacrificed to obtain sera. Animal experiments were approved by the St. Jude Children’s Research Hospital Animal Care and Use Committee.

### Serum neutralization assay

Human or ferret serum was used in microneutralization assays using MDCK cells as previously described [10]. The neutralizing antibody titer was calculated as the highest serum dilution that eliminated virus cytopathic effects in 50% of the wells.

### Statistical analysis

Serology data were analyzed with a 2-way ANOVA followed by pairwise multiple comparisons or by 1-way ANOVA or students t-test. Fold change in antibody titers between different virus strains and across different sites were analyzed by Chi-square analysis. Virus replication kinetics were analyzed using a repeated measures 2-way ANOVA. Clinical data in tables 1-3 were analyzed by Chi-square analysis.

**Table 1.** Patient characteristics at the two geographic locations, Baltimore and Taipei.

|  | Baltimore |  |  | Taipei |  |  |
| --- | --- | --- | --- | --- | --- | --- |
|  | ILI+, IAV+ | ILI+, IAV- | p-value | ILI+, IAV+ | ILI+, IAV- | p-value |
| **Age** |  |  |  |  |  |  |
| 18-49 | 70.6% (60/85) | 64.7% (108/167) | 0.3973 | 62.2% (92/148) | 68.0% (66/97) | 0.501 |
| 50-64 | 23.5% (20/85) | 26.9% (45/167) | 0.6483 | 25.0% (37/148) | 24.7% (24/97) | 1 |
| 65+ | 5.9% (5/85) | 8.4% (14/167) | 0.6166 | 12.8% (19/148) | 7.2% (7/97) | 0.2048 |
| **Race** |  |  |  |  |  |  |
| White | 23.5% (20/85) | 40.1% (67/167) | 0.0114 | 0% (0/148) | 0% (0/97) | 1 |
| Black or African American | 67.1% (57/85) | 54.5% (91/167) | 0.0594 | 0% (0/148) | 0% (0/97) | 1 |
| Asian | 0% (0/85) | 1% (1/167) | 1 | 100% (148/148) | 100% (97/97) | 1 |
| Other | 9.4% (8/85) | 4.8% (8/167) | 0.1765 | 0% (0/148) | 0% (0/97) | 1 |
| **Gender** |  |  |  |  |  |  |
| Female | 52.9% (45/85) | 57.5% (96/167) | 0.5051 | 65.5% (97/148) | 52.6% (51/97) | 0.0461 |
| Male | 47.1% (40/85) | 42.5% (71/167) |  | 34.5% (51/148) | 47.4% (46/97) |  |
| **Vaccination** |  |  |  |  |  |  |
| Vaccinated | 29.8% (25/84) | 50.3% (82/163) | 0.0028 | 6.1% (9/148) | 12.4% (12/97) | 0.1037 |
| **Co-morbidities** |  |  |  |  |  |  |
| None | 35.3% (30/85) | 19.8% (33/167) | 0.0089 | 54.7% (81/148) | 47.4% (46/97) | 0.2392 |
| One | 29.4% (25/85) | 29.9% (50/167) | 1 | 20% (29/148) | 29% (28/97) |  |
| Multiple | 35.3% (30/85) | 50.3% (84/167) | 0.0319 | 25.7% (38/148) | 23.7% (23/97) |  |
| **Pregnancy** |  |  |  |  |  |  |
|  | 6.7% (3/45) | 3.2% (3/95) | 0.3861 | 5.2% (5/97) | 5.9% (3/51) | 1 |
| **Disease severity** |  |  |  |  |  |  |
| Oxygen supplementation | 17.6% (15/85) | 28.7% (48/167) | 0.0648 | 23.0% (34/148) | 17.5% (17/97) | 0.3374 |
| Intubation | 0% (0/84) | 0% (0/167) | 1 | 1% (1/148) | 3% (3/97) | 0.3033 |
| Inpatient | 29.4% (25/85) | 44.3% (74/167) | 0.0288 | 31% (46/148) | 44% (43/97) | 0.0584 |
| Length of stay | 3 (1-12) | 2 (1-21) | 0.8824 | 5 (1-55) | 6 (1-59) | 0.8476 |
| ICU | 8.0% (2/25) | 0% (0/74) | 0.0618 | 4.3% (2/46) | 7.0% (3/43) | 0.6729 |
| Pneumonia | 18% (15/85) | 28% (46/167) | 0.1528 | 41% (60/148) | 40% (39/97) | 1 |
| Death | 0% (0/85) | 0% (0/167) | 1 | 4% (6/148) | 0% (0/97) | 0.0839 |

For multivariate analyses, the antibody titers were log transformed. All variables with *p* < 0.2 and beta coefficient > 0.1 in the bivariate linear regression were evaluated with multivariate linear regression with Allen-Cady backward selection procedure, with priority determined by clinical relevance and existing literature support. We adopted the Akaike’s Information Criteria and the Chi-square goodness of fit test to evaluate the model fitness. Interaction check was performed to evaluate the potential effect modification between vaccination, antibody titers, and significant risk factors. Because of the relative small sample size, we further performed propensity score weighting modeling, in order to simultaneously adjust multiple confounders. Confounders considered to influence the causal relationship between vaccination and antibody titers were modeled to generate the propensity of vaccination, with the propensity score used as the weight to balance the potential confounding and evaluate the average treatment effect. Mean differences were considered statistically significant if *p*<0.05.

## Results

Influenza surveillance was performed in the ED at JHMI and at CGMH from November 1, 2015 (week 44 of 2015) to April 30, 2016 (week 17 of 2016). The basic demographic and disease characteristics of patients reporting with ILI that were either confirmed to be infected (IAV+) or not (IAV-) in Baltimore (n=252) and in Taipei (n=245) are summarized in **Table 1**. In Baltimore, there was a racial difference, in which more Caucasian/white patients with ILI were IAV- (40.1%) than IAV+ (23.5%). For IAV vaccination in Baltimore, 50.3% of IAV-patients received the 2015-16 seasonal vaccine compared to 29.8% in the IAV+ group. The Baltimore IAV- population had significantly more patients with multiple co-morbidities (50.3%) than the IAV+ population (35.3%). There was also a greater number of IAV- patients with ILI admitted to JHMI hospitals (44.3%) than IAV+ patients (29.4%). In contrast, fewer differences were observed between the IAV+ and IAV- individuals with ILI from the Taipei population. Greater numbers of females as compared to males were present in the IAV+ group (65.5%) than the IAV- group (52.6%) in Taipei.

Enrollment weeks for IAV+ individuals were plotted (**Figure 1A and 1B**). Influenza A virus activity was lower and peaked later (mid-March; **Figure 1A**) in the Baltimore population and was higher and peaked earlier (January-February; **Figure 1B**) in the Taipei population which was representative of the overall IAV activity in each country [6, 7].

**Figure 1.**
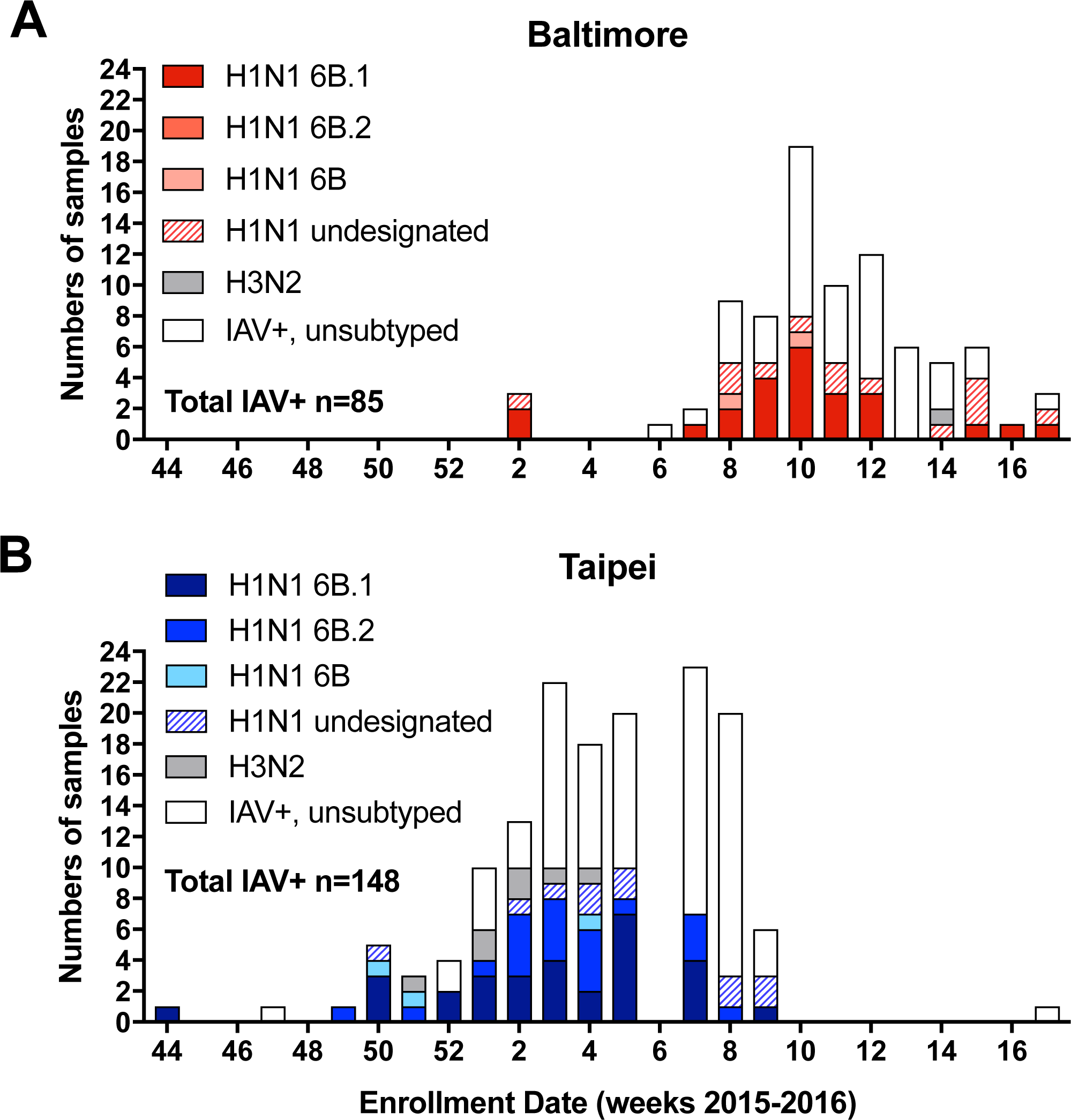
Numbers of Influenza A virus isolates defined by clades with enrollment dates in the two surveillance sites, Baltimore (JHMI) and Taipei (CGMH). Samples collected from patients with influenza-like illness (ILI) between week 44 2015 (starting Nov 2015) and week 16 2016 (ending April 2016) were tested for influenza A viruses (IAVs) and the IAV+ samples were subtyped and clade-defined by deep sequencing. (A) JHMI, (B) CGMH. Enrollments at week 6 of 2016 in Taipei were paused due to the Chinese New Year holidays.

Among the 233 IAV+ cases across the surveillance sites, 111 NW or NP swabs were processed for sequencing. H1N1 was the predominant IAV subtype circulating globally [6, 17, 18] and in both Baltimore and Taipei (**Figure 1A** and **1B**). One-hundred and three of the isolates (92.8%) were identified as H1N1 viruses and only 8 (7.2%) were H3N2 viruses. Seventy-nine of the 103 H1N1 clinical virus isolates yielded sufficient coverage of the HA segment and all belonged to clade 6 subgroup B (clade 6B), 6B.1 or 6b.2 [19], with the latter two clades newly emerged in this season [20]. Phylogenetic analysis of the HA gene sequences from patients in Baltimore (red) or Taipei (blue) classified most isolates as clade 6B.1. Clade 6B.2 viruses made up a significant number of the isolates in Taipei, but were absent in Baltimore (**Figure 2A and 2B**).

**Figure 2.**
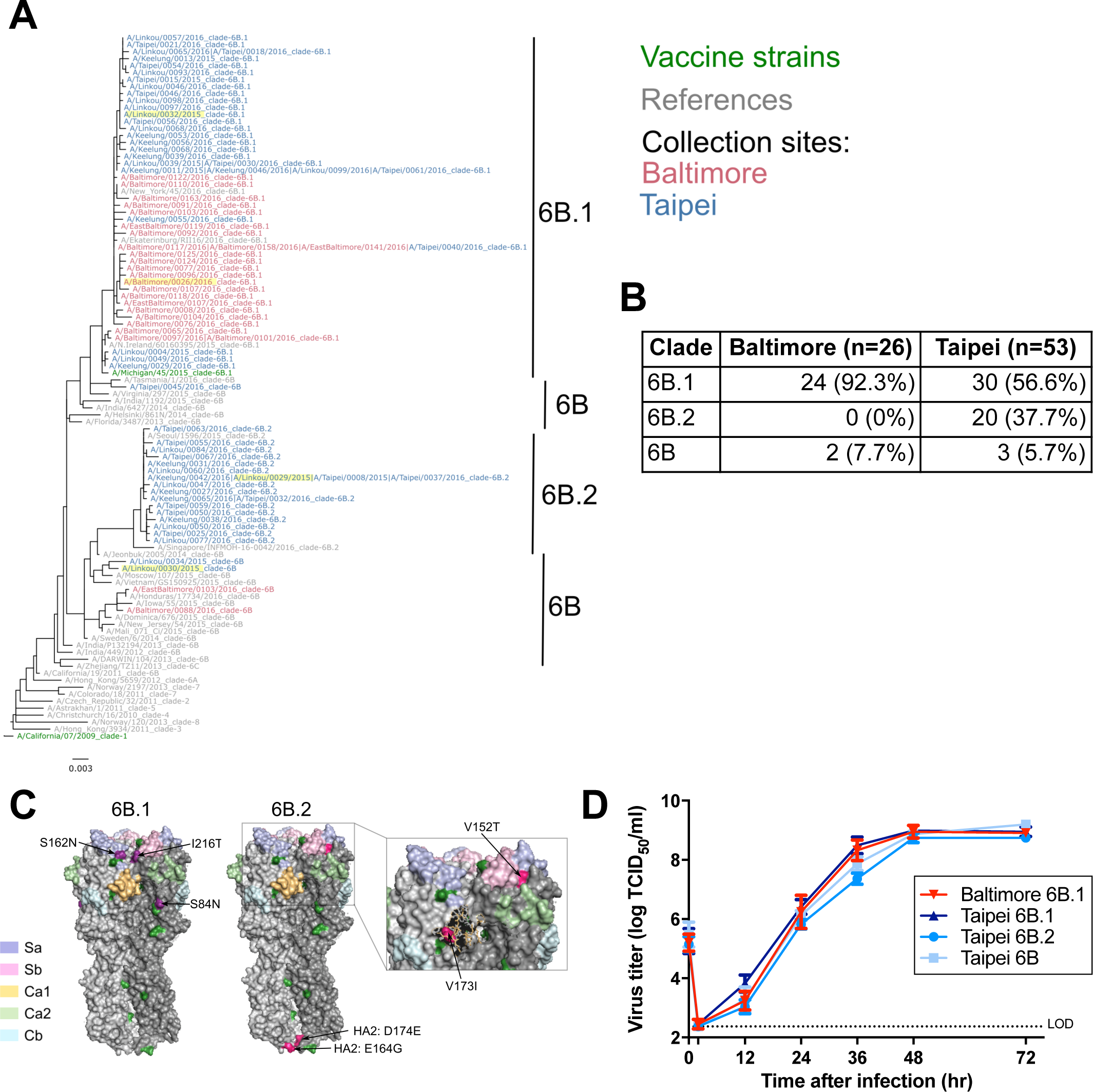
H1N1 HA phylogenetic analysis from clinical virus isolates. Clinical nasal pharyngeal (NP) swabs or nasal washes (NW) of IAV+ patients from Baltimore (JHMI) and Taipei (CGMH) were used to sequence the HA gene segment by deep sequencing. (A) HA phylogenetic tree representing the 2015-16 H1N1 viruses from individuals at Baltimore (red) and Taipei (blue) along with the 2015-16 vaccine strain A/California/7/2009 (green) and the 2017-18 vaccine strain A/Michigan/45/2015 (green). Representative isolates for serology assays and growth analysis are highlighted in yellow. (B) Numbers of clinical virus isolates in clades 6B.1, 6B.2 and 6B were identified. (C) HA protein structure of the clade 6B.1 and 6B.2 viruses are shown on a H1 HA crystal structure of H1N1 virus (PDB:3LZG) using PyMOL. Antigenic sites Sa, Sb, Ca1, Ca2, and Cb are indicated [46]. Amino acids are used to defined 6B.1 (purple) and 6B.2 (pink) and other amino acid differences compared with A/California/7/2009 (dark green) are indicated. Enlarged 6B.2 head shows that aa173 (in pick spheres) is not on the surface of HA, and is under site Ca1 (in sticks). (D) Viruses isolated from individuals at both Baltimore and Taipei were propagated on hNECs. Multistep growth curves were performed on hNECs at 32°C with the representative viral isolates: clade 6B.1 virus from Baltimore and of clade 6B.1, 6B.2, and 6B viruses from Taipei. Representative data from 2 independent experiments are shown. Dotted line indicates limit of detection.

A comparison of the patient populations of Baltimore and Taipei that were infected with H1N1 viruses revealed that these populations were similar in terms of demographics and disease severity (**Table 2**), with expected differences in race observed between sites. More patients with no known co-morbidities for severe influenza were infected in the Taipei population compared to Baltimore. Vaccine coverage was higher in the Baltimore population (31.6%) than the Taipei population (4.7%). These data show stronger influenza activity and H1N1 infection in Taipei compared to Baltimore. Outside of race, there were no differences in the clinical or demographic characteristics of patients infected with clade 6B.1 viruses in Baltimore when compared to those infected with 6B.1 viruses in Taipei (**Supplementary Table 2**). Among patients infected with clade 6B.1 viruses at the two sites, there were no significant demographic differences. Likewise, there were no differences in Taipei patients infected with 6B.1 compared to 6B.2 viruses (**Supplementary Table 2**), indicating that the disease incidence and severity differences between the Baltimore and Taipei sites were not driven by any specific demographic or clinical factor.

**Table 2.**
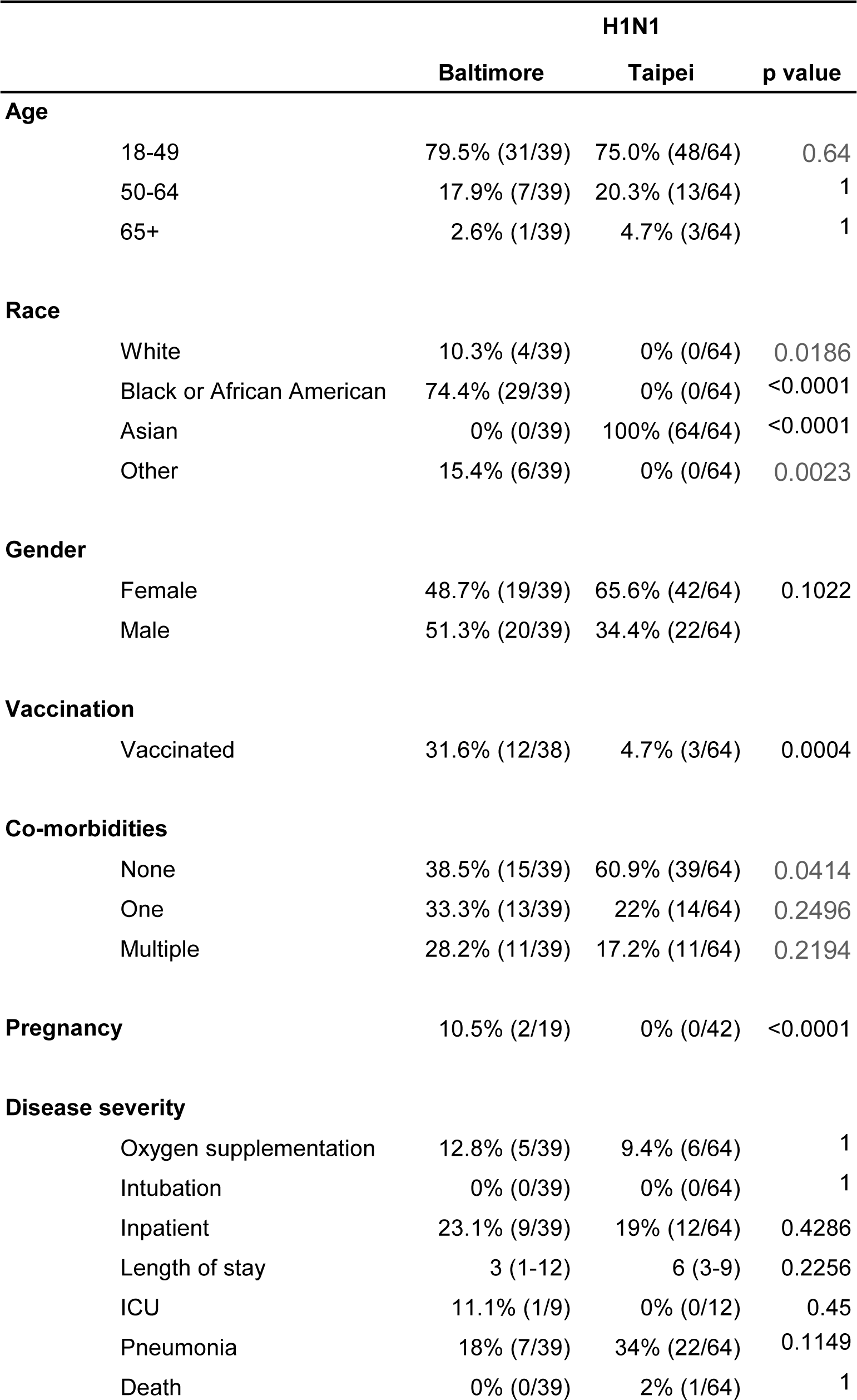
Clinical and demographic characteristics of H1N1 IAV-infected patients at Baltimore and Taipei.

|  |  | H1N1 |  |  |
| --- | --- | --- | --- | --- |
|  |  | Baltimore | Taipei | p value |
| **Age** |  |  |  |  |
|  | 18-49 | 79.5% (31/39) | 75.0% (48/64) | 0.64 |
|  | 50-64 | 17.9% (7/39) | 20.3% (13/64) | 1 |
|  | 65+ | 2.6% (1/39) | 4.7% (3/64) | 1 |
| **Race** |  |  |  |  |
|  | White | 10.3% (4/39) | 0% (0/64) | 0.0186 |
|  | Black or African American | 74.4% (29/39) | 0% (0/64) | <0.0001 |
|  | Asian | 0% (0/39) | 100% (64/64) | <0.0001 |
|  | Other | 15.4% (6/39) | 0% (0/64) | 0.0023 |
| **Gender** |  |  |  |  |
|  | Female | 48.7% (19/39) | 65.6% (42/64) | 0.1022 |
|  | Male | 51.3% (20/39) | 34.4% (22/64) |  |
| **Vaccination** |  |  |  |  |
|  | Vaccinated | 31.6% (12/38) | 4.7% (3/64) | 0.0004 |
| **Co-morbidities** |  |  |  |  |
|  | None | 38.5% (15/39) | 60.9% (39/64) | 0.0414 |
|  | One | 33.3% (13/39) | 22% (14/64) | 0.2496 |
|  | Multiple | 28.2% (11/39) | 17.2% (11/64) | 0.2194 |
| **Pregnancy** |  | 10.5% (2/19) | 0% (0/42) | <0.0001 |
| **Disease severity** |  |  |  |  |
|  | Oxygen supplementation | 12.8% (5/39) | 9.4% (6/64) | 1 |
|  | Intubation | 0% (0/39) | 0% (0/64) | 1 |
|  | Inpatient | 23.1% (9/39) | 19% (12/64) | 0.4286 |
|  | Length of stay | 3 (1-12) | 6 (3-9) | 0.2256 |
|  | ICU | 11.1% (1/9) | 0% (0/12) | 0.45 |
|  | Pneumonia | 18% (7/39) | 34% (22/64) | 0.1149 |
|  | Death | 0% (0/39) | 2% (1/64) | 1 |

The amino acid mutations in the HA protein of circulating virus strains were mapped on the H1 HA protein crystal structure (**Figure 2C**). Clade 6B.1 viruses were characterized by mutations S84N, S162N, and I216T (purple), with an amino acid residue 162 on the Sa antigenic site (blue). Amino acid 216, along the 220 loop of the receptor-binding domain was adjacent to the Sb antigenic site (pink). The I216T mutation may be a compensatory mutation as a result of the S162N mutation [21]. Clade 6B.2 viruses contained two HA1 mutations: V152T, adjacent to the Sa and Sb antigenic sites and V173I, near the Ca1 antigenic site (yellow). In addition to the mutations in HA1, 6B.2 viruses contained two mutations in HA2: E164G and D174E which were both on the surface of the HA stalk domain (**Figure 2C**). When compared to the vaccine strain A/Cal/7/09, both clade 6B.1 and 6B.2 viruses had 11 additional amino acid mutations (P83S, D97N, K163Q, S185T, S203T, A256T, K283E, I321V and HA2: E47K, S124N, and E172K), many of which were located on or near antigenic sites.

The replication efficiency of 6B.1 and 6B.2 clinical virus isolates was assessed using hNEC cultures. No significant differences in virus replication kinetics were detected (**Figure 2D**) with viruses isolated from the two surveillance sites.

Influenza-specific antibodies are considered the primary correlate of protection against influenza infection in humans [22]. Neutralizing antibody responses to the clade 6B.1 virus were greater in patients from Baltimore than Taipei in serum collected at the time of enrollment (**Figure 3A**). The higher titer of anti-clade 6B.1 antibodies, but not antibodies to the vaccine strain, in the Baltimore population indicated higher titers of antibodies to circulating viruses that do not cross react with the vaccine strain and suggested that human antibodies can differentiate between the H1N1 circulating and vaccine strains.

**Figure 3.**
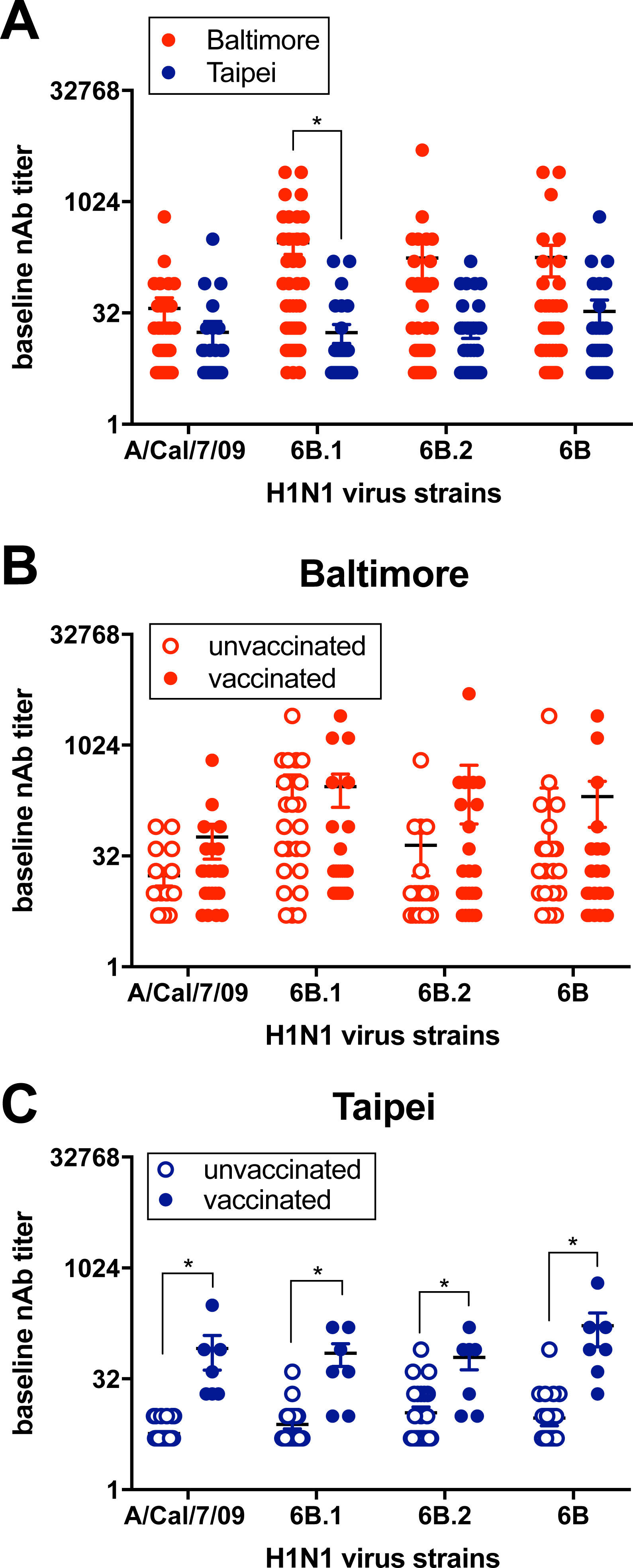
Baseline human serum neutralizing antibody titers to vaccine and circulating strains of H1N1. Human serum was collected at the time of enrollment and neutralizing antibody (nAb) titers to the vaccine strain A/Cal/7/09, and representative circulating strains 6B.1, 6B.2, and 6B were determined by neutralization assay. (A) Baseline neutralizing antibody titers from individuals at Taipei (CGMH) (n=46) and Baltimore (JHMI) (n=45), (B) vaccinated individuals (n=22) and unvaccinated individuals (n=23) from Baltimore, and (C) vaccinated individuals (n=7) and unvaccinated individuals (n=39) from Taipei.

We compared individuals that received the 2015-16 seasonal influenza vaccine with those that did not across the two surveillance sites. Although receipt of the seasonal vaccine did not affect neutralizing antibody responses to the vaccine or circulating viruses in patients from Baltimore (**Figure 3B**), neutralizing antibody responses from Taipei patients were significantly greater in vaccinated compared to unvaccinated patients (**Figure 3C**). We conclude that geographic location and presumably the history of exposure to circulating and vaccine strains of influenza impact the antibody response induced by vaccination.

The H1N1 component of the seasonal influenza vaccine was updated for the 2017-18 influenza season from A/Cal/7/09 to clade 6B.1 A/Mich/45/15 [23], so we hypothesized that the baseline neutralizing antibody responses to this virus would be similar to responses to the 2015-16 circulating clade 6B.1 viruses. Both vaccinated and unvaccinated patients from Baltimore had cross-reactive neutralizing antibody responses to the A/Mich/45/15 vaccine virus, whereas in Taipei only vaccinated individuals had cross-reactive neutralizing antibody responses as compared to unvaccinated individuals (**Figure 4A**), indicating that vaccination could have regional differences in the induction of cross-reactive antibody responses.

**Figure 4.**
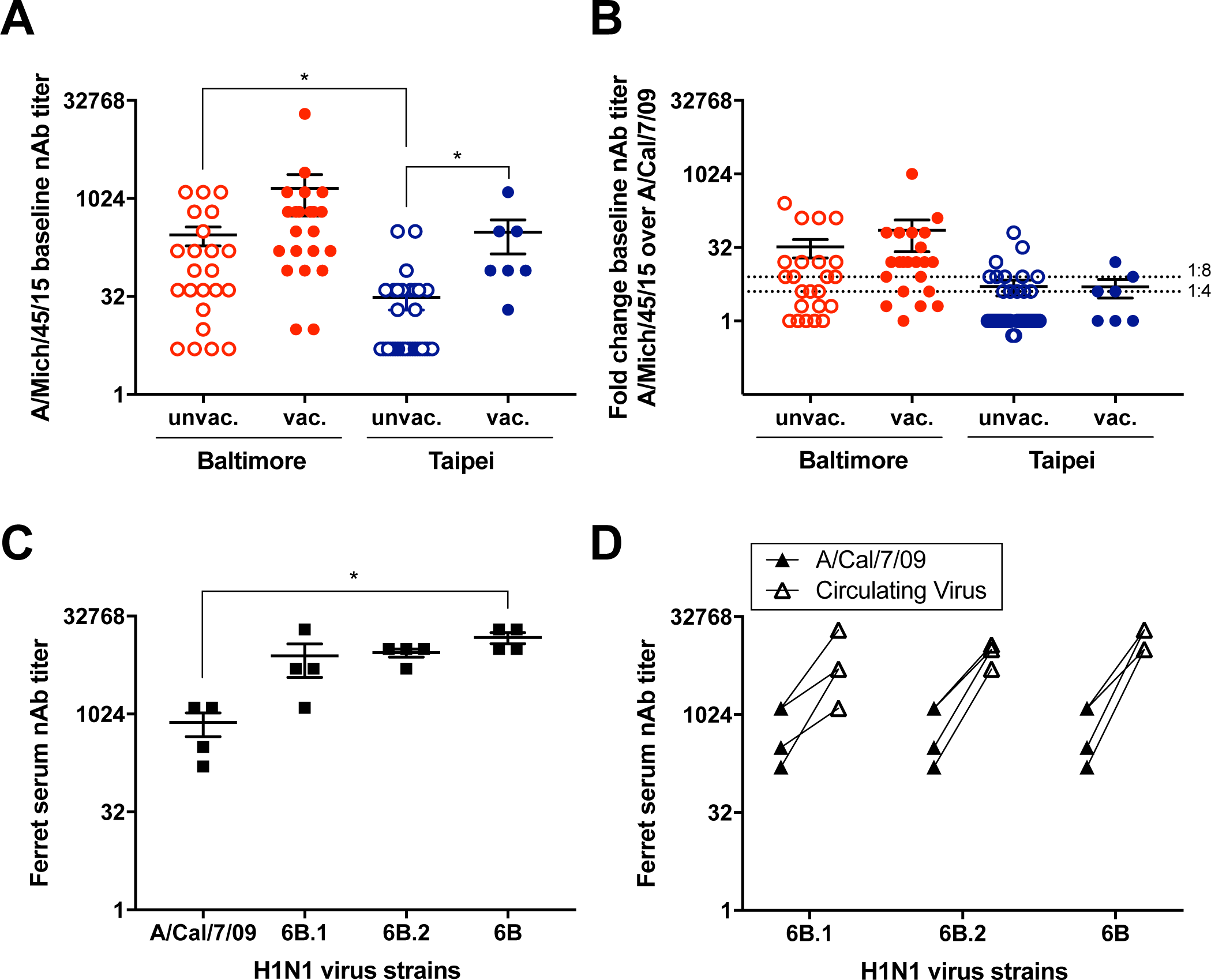
Human and ferret serum reactivity to H1N1 vaccines strains. Human serum was collected at the time of enrollment and neutralizing antibody (nAb) titers to the H1N1 vaccine strains A/Cal/7/09 and A/Mich/45/15 were determined by neutralization assay. (A) nAb titers to A/Mich/45/15 from vaccinated (n=22) and unvaccinated (n=23) individuals from Baltimore (JHMI) and vaccinated (n=7) and unvaccinated (n=39) individuals from Taipei (CGMH). To compare the baseline nAb titers between vaccine strains A/Cal/7/09 and A/Mich/45/15, individual baseline nAb titers to A/Mich/45/15 and A/Cal/7/09 in vaccinated and unvaccinated individuals from Baltimore and Taipei were plotted (B). Male ferrets were infected with either A/Baltimore/0026/2016 (n=2) or A/Linkou/0029/2015 (n=2) and 21 days later serum was collected to determine the neutralizing antibody responses to A/Cal/7/09 and 6B.1, 6B.2, and 6B circulating viruses (C). The individual ferret serum neutralizing antibody responses to the A/Cal/7/09 virus as compared to the circulating strains (6B.1, 6B.2, 6B) was then plotted (D).

We analyzed the baseline neutralizing antibody titers to the A/Cal/7/09 and the A/Mich/45/15 vaccine strain. The serum cross-reactivity to the A/Mich/45/15 vaccine virus as compared to the A/Cal/7/09 vaccine virus was greater in Baltimore compared Taipei using a ≥ 4 fold increase in titer as a cutoff (**Figure 4B**) which was driven by differences among unvaccinated patients at the two surveillance sites. Using a more stringent cutoff of ≥ 8 fold increase in titer, there was still greater recognition of the A/Mich/45/15 vaccine virus among patients from Baltimore as compared to those from Taipei, with both unvaccinated and vaccinated populations driving the cross-site differences (**Figure 4B**), indicating there are regional differences in the recognition of vaccine strains and the circulating 6B.1 viruses (**Figure 3A** and **4A**).

The antigenic characterization of IAV isolates is traditionally measured in HAI assays using strain specific, post-infection ferret antisera [24] and HAI assays showed ferret sera raised to circulating reference 6B.1 and 6B.2 viruses cross-reacted well with the A/Cal/7/09 vaccine virus [19]. Using sera from ferrets infected with the circulating H1N1 virus strains from Baltimore and Taipei, there were greater neutralizing antibody responses to the clade 6B virus as compared with the A/Cal/7/09 vaccine virus (**Figure 4C**). All ferrets had a ≥ 4 fold increase in titer specific to each of the circulating virus strains as compared to the A/Cal/7/09 vaccine strain (**Figure 4D**). Our data showed low cross-reactivity between the ferret sera raised to circulating H1N1 strains and the A/Cal/7/09 vaccine strain, indicating potential antigenic differences between the circulating and vaccine strains, and supporting the recommendation to update the H1N1 component of the seasonal influenza vaccine.

Using an Allen-Cady modified stepwise multivariable regression model to control for confounding variables revealed increased age was consistently associated with lower titers (**Table 3**). In contrast, a significant association between age and neutralizing antibody titers in our univariate analysis was not observed (**Supplementary Figure 1**). Other factors that were associated with lower titers in different clades included male gender (A/Cal/7/09, 6B.1), steroid usage (6B), higher BMI (6B.2), being Asian (6B.1), and some socioeconomic factors. Although we did not observe gender differences in baseline neutralizing antibody titers in our univariate analysis (**Supplementary Figure 2**), previous studies have established that males tend to have significantly lower neutralizing antibody titers to influenza as compared with females [25]. Receipt of the influenza vaccine was associated with higher baseline neutralizing antibody titers to the A/Cal/7/09 vaccine strain virus and the clade 6B and 6B.2 circulating viruses. In a propensity score weighting model, receipt of the influenza vaccine resulted in higher neutralizing antibody titers to all viruses except the clade 6B.1 virus. In the multivariable regression, when the variable “Asian” was removed, vaccination was still significantly associated with higher baseline neutralizing antibody titers, indicating that there could be a confounding effect between being Asian and receipt of the influenza vaccine. In the effect modification multivariable regression analysis, we found that vaccinated patients from Taipei had consistently higher antibody titers across different clades, except 6B.2 (**Table 3**). The observation that receipt of the influenza vaccine was associated with higher baseline neutralizing antibody titers to all viruses except the clade 6B.1 virus further confirms the recommendation to update the H1N1 vaccine virus to A/Mich/45/15, which is a clade 6B.1 virus.

**Table 3.**
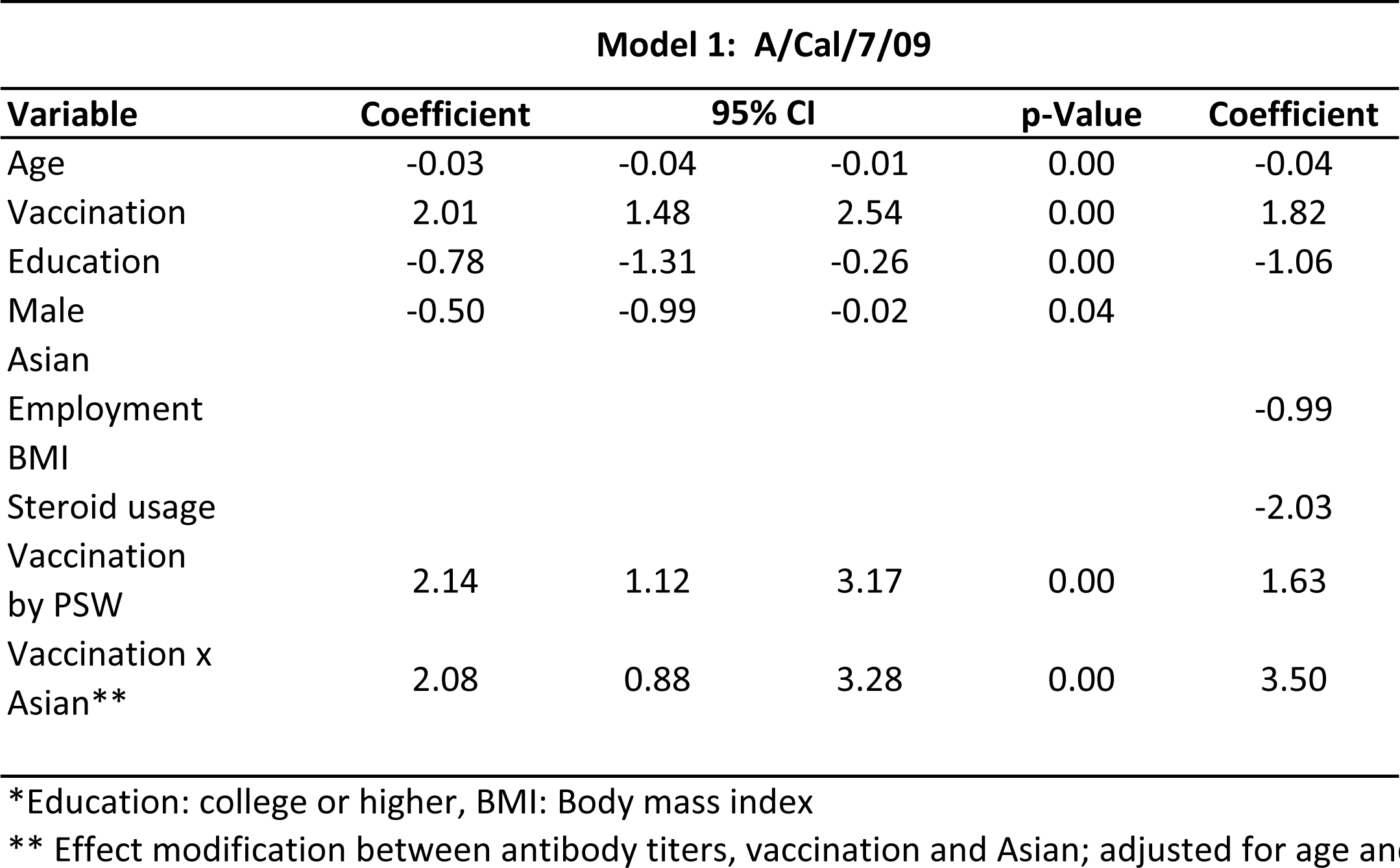
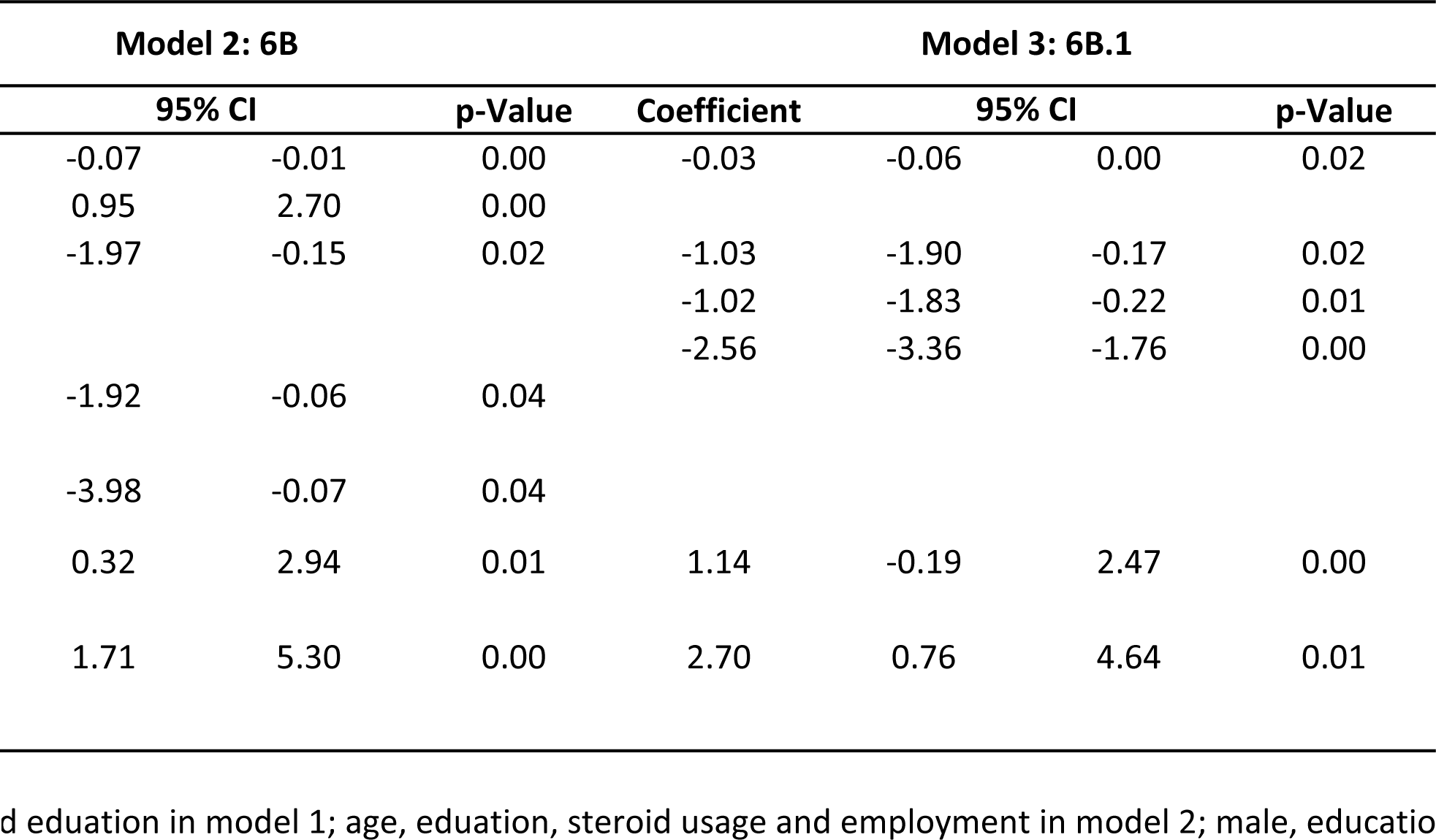
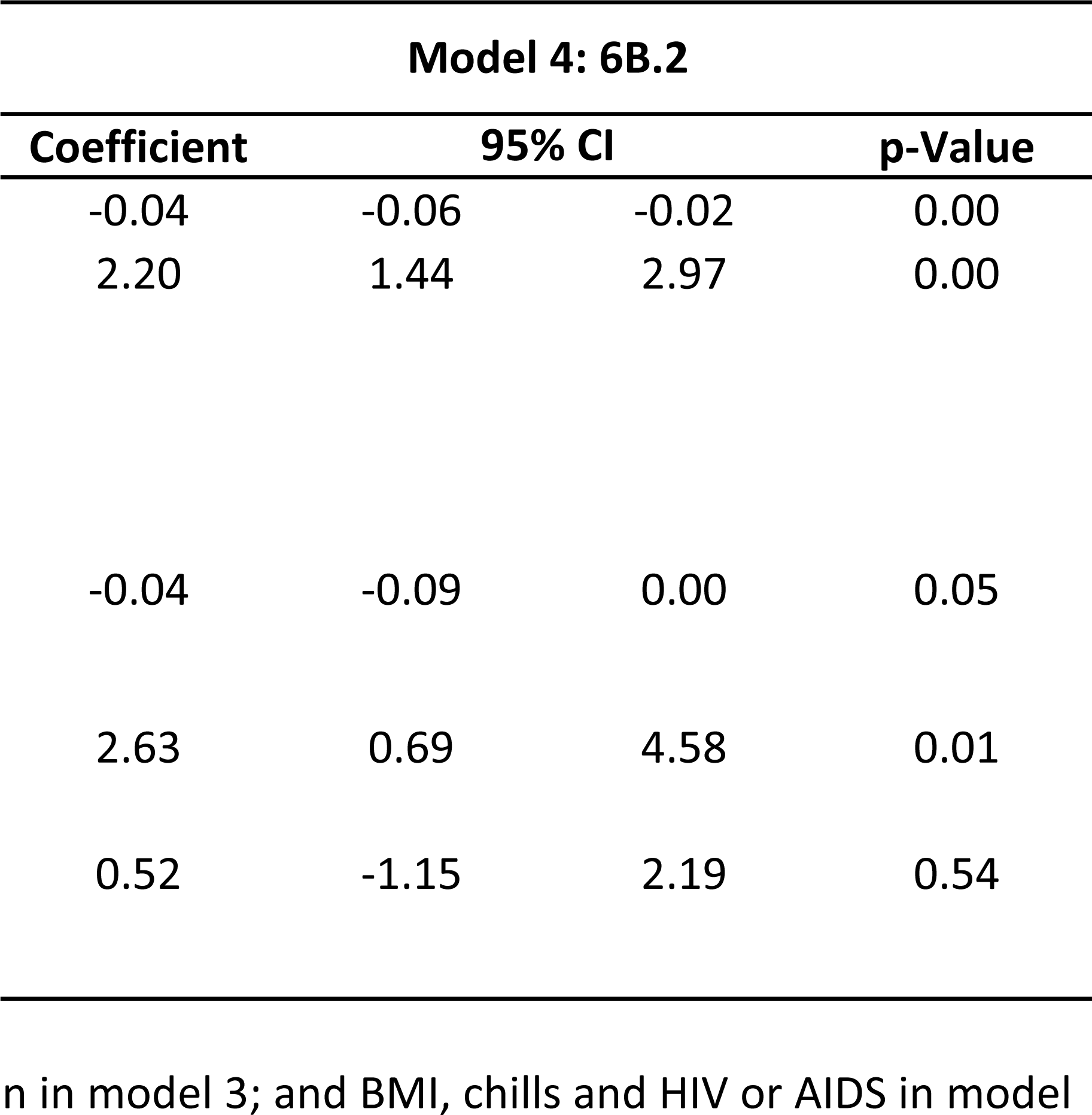
Allen-Cady multivariate analysis.

## Discussion

This study illustrates differences in influenza activity, pre-existing influenza immunity, and virus strain circulation in the 2015-16 influenza season across surveillance sites located in Baltimore, Maryland and Taipei, Taiwan. We conclude that regional factors as well as host demographic variables, including age, gender, and BMI, may play important roles in modulating receipt of seasonal influenza vaccines, vaccine-induced immunity, and severity of influenza disease.

The 2015-16 influenza season was characterized by increased H1N1 influenza cases in Taipei compared to Baltimore, with differences in the HA clades that circulated at each site, consistent with previous reports [26-28]. Clade 6B.2 viruses circulated in primarily in Asia but never became a common clade globally. While specific amino acid changes near antigenic sites are present in clade 6B.2 viruses, they appear to be antigenically indistinguishable from clade 6B.1 viruses using human and ferret serum. Both H1N1 clades had similar replication kinetics in hNEC cultures, indicating there were no inherent differences in virus fitness between the two clades. Since the 2016-17 season, only H1N1 6B.1 viruses have been detected and have evolved to different clade 6B.1 subclades which are continuing circulation worldwide [23, 29-32].

The demographic characteristics of the Baltimore and Taipei sites were similar, except for race. It is difficult to ascertain the contribution of race to influenza susceptibility given these differences. There was a significantly larger number of influenza cases in individuals with no known influenza co-morbidities in Taipei compared to Baltimore, indicating a greater degree of susceptibility in the population in Taipei, which is supported by the overall lower vaccine rate and levels of neutralizing antibodies to circulating human H1N1 present in that population. The overall lower antibody titers to 6B.1 viruses in Taipei were surprising given that antigenically related viruses have been circulating globally since 2009. Vaccination, however, did not affect neutralizing antibody titers in patients from Baltimore, perhaps due to higher levels of preexisting H1N1 immunity in that population. These data suggest that regional differences in influenza vaccine uptake and immune responses to vaccination are present and can impact the overall susceptibility of the population to influenza virus infection.

Global 2015-16 influenza surveillance in most countries indicated the circulating H1N1 viruses were antigenically similar to the vaccine A/Cal/7/09 [6-9, 17, 33, 34]. In our study, antigenic differences between the H1N1 vaccine strain and circulating viruses in 2015-16 were detected using human serum and serum from ferrets infected with circulating H1N1 viruses in neutralization assays. Consistent with those observations, higher neutralizing antibody responses to the A/Mich/45/15 vaccine strain compared to the A/Cal/7/09 vaccine strain were detected. Studies also showed that humans elicited differential antibodies recognizing cell-derived or egg-adapted vaccines which was driven by the egg-adaption mutation Q226R (which is Q223R in H1 numbering) of A/Cal/7/09, thus causing some individuals to better recognize the vaccine strain in HAI assays [35-37]. However, we did not see significant differences in neutralization titers of 6B.1 circulating virus compared to the A/Mich/45/15 vaccine containing Q223R (Figure 3A 6B.1 and Figure 4A). Even though ferret antiserum were unable to distinguish between A/Cal/7/09 and A/Mich/45/15, the WHO changed the H1N1 vaccine component to A/Mich/45/15 in the 2017 Southern hemisphere vaccine and subsequently in the 2017-18 Northern hemisphere because circulating viruses were poorly recognized by some post-vaccination adult human serum [38, 39]. Our use of human sera and virus neutralization assays strongly indicate antigenic differences between the vaccine and circulating strains.

We did not observe any relationship between neutralizing antibody titers and specific age-ranges [40-43], but this is most likely due to small sample sizes in most age groups. Although more females presented as IAV+, at least in Taipei, male gender, steroid use, and higher BMI correlated with lower neutralizing titers, as these are associated with an impaired humoral immune response [44, 45].

There are limitations of this study. Our acute influenza detection protocol does not include pediatric patients so we are unable to assess preexisting immunity and case numbers in age groups <18 years of age. Because only one surveillance site is used in each country, we are unable to capture any regional differences within the United States or Taiwan. Finally, the limited number of patients enrolled in our studies prevents us from assessing the significance of a number of clinical and demographic factors on influenza susceptibility and disease severity.

Although seasonal influenza epidemics are global, regional differences in human populations can impact the number and severity of influenza cases most likely through different exposure histories to circulating and vaccine strains. Vaccine efficacy and preexisting immunity to vaccine and circulating strains should be characterized at the local, national, and global levels to achieve accurate and complete assessments of the factors that impact influenza in any year.

## Data Availability

All data is available for analysis by request.

## Acknowledgements

We would like to thank the clinical coordinators at JHMI and CGMH who were responsible for the data and sample collection. We also thank Stacey Schultz-Cherry, Victoria Meliopoulos, and the St. Jude Center of Excellence in Influenza Research and Surveillance (CEIRS) for providing the ferret antisera. We thank Deena Blumenkrantz and Robert Stenzel for providing technical assistance in growing influenza viruses and Dr. Andrew Lane for providing hNEC cultures.

---

**Supplementary Figure 1.**
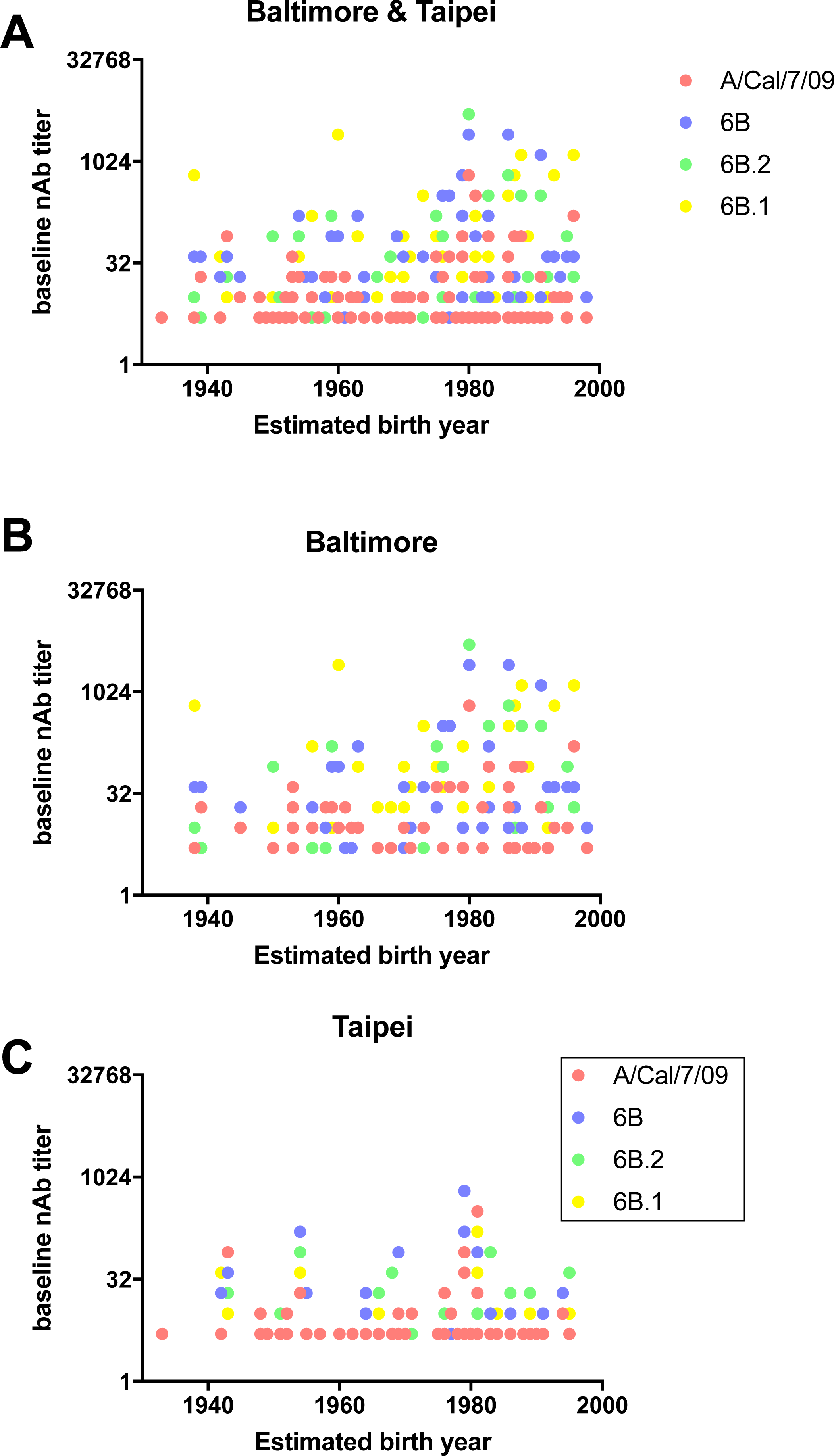
Univariate analysis of baseline neutralizing antibody titers based on estimated birth year. Each individual’s baseline neutralizing antibody titers to the A/Cal/7/09, 6B, 6B.2, and 6B.1 viruses were plotted against their estimated birth year. Individuals from both sites (A), individuals from Baltimore (JHMI) (B) and individuals from Taipei (CGMH) (C) are represented. Correlation analysis was performed and no significant correlations were observed.

**Supplementary Figure 2.**
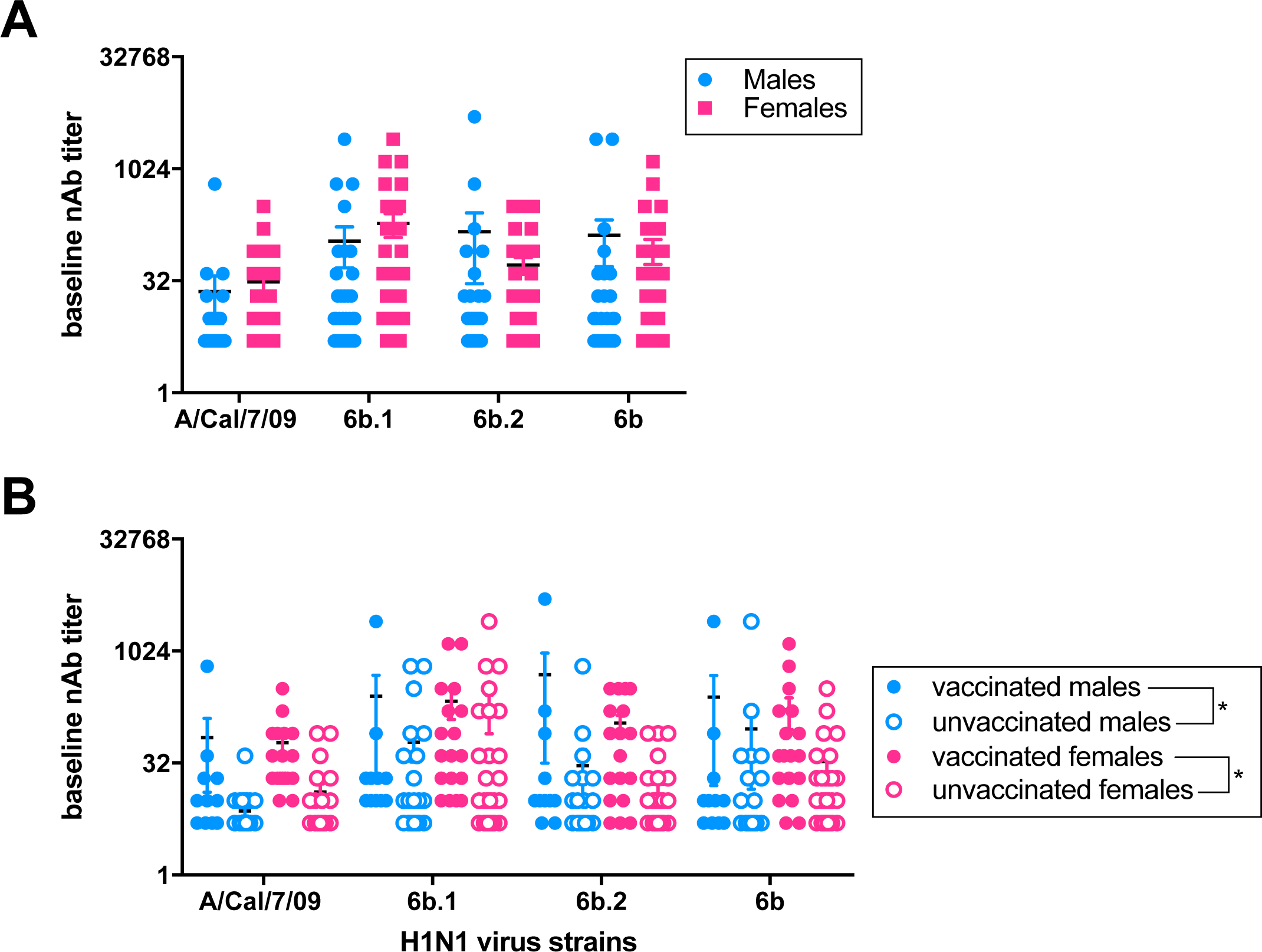
Univariate analysis of gender differences in baseline neutralizing antibody titers. Baseline neutralizing antibody titers to the A/Cal/7/09, 6B, 6B.2, and 6B.1 viruses were evaluated in individuals from Baltimore (JHMI) and Taipei (CGMH) and data were stratified by gender. Data were first stratified by gender alone (A) and then further stratified based on both gender and vaccine status (B).

**Supplementary Table 1:**
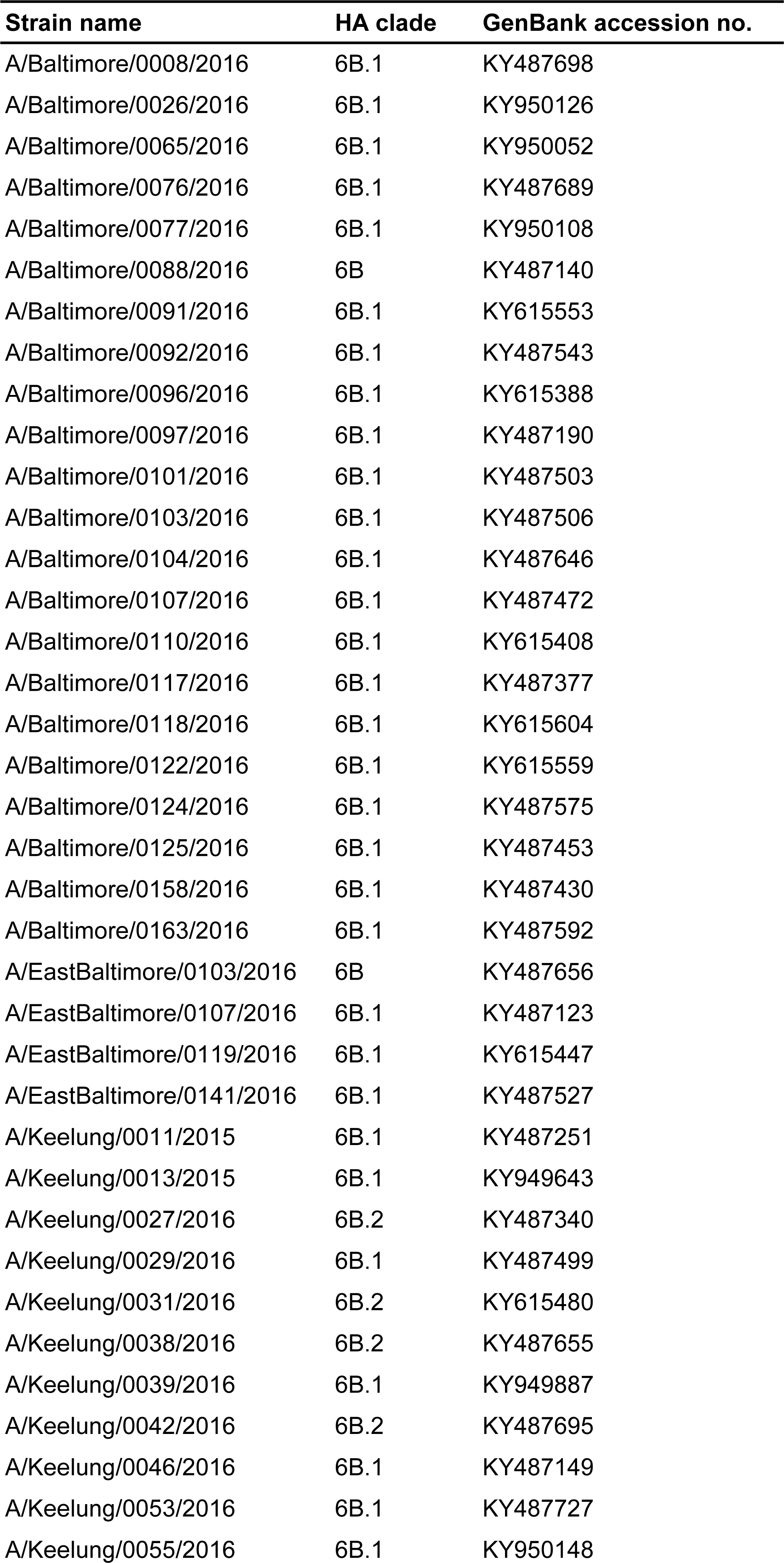
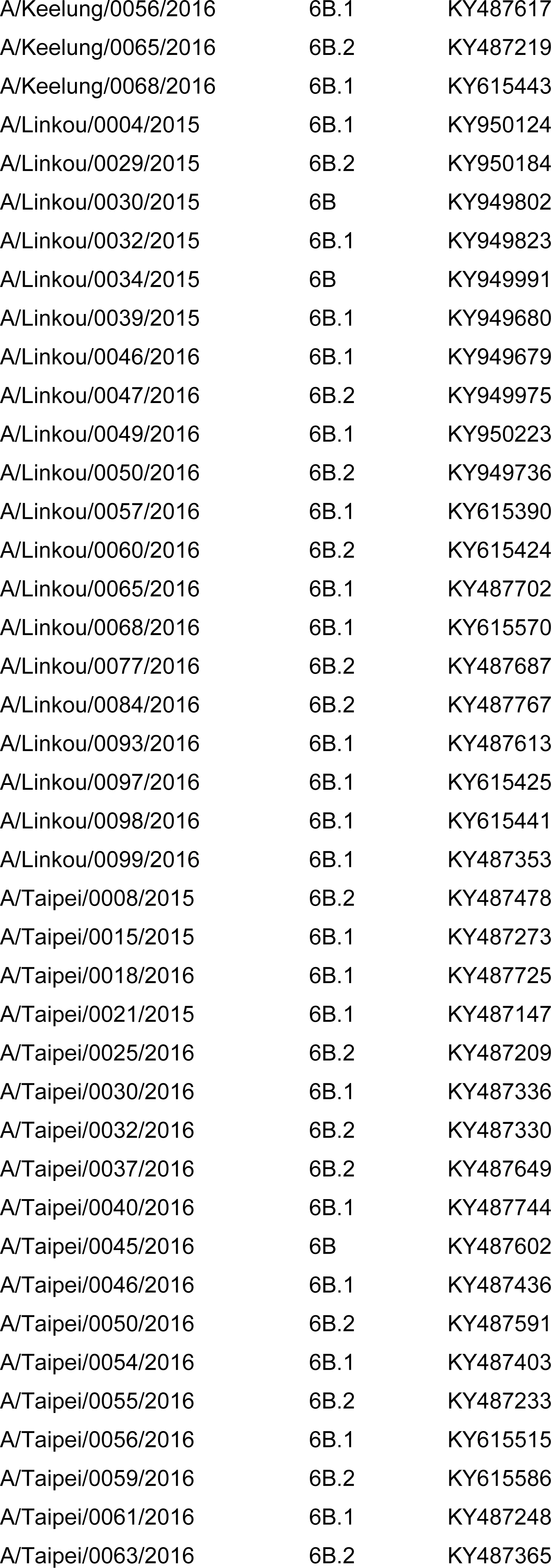
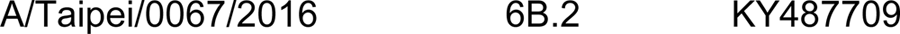
Influenza A virus strains and assoicated GenBank accession numbers.

**Supplementary Table 2.**
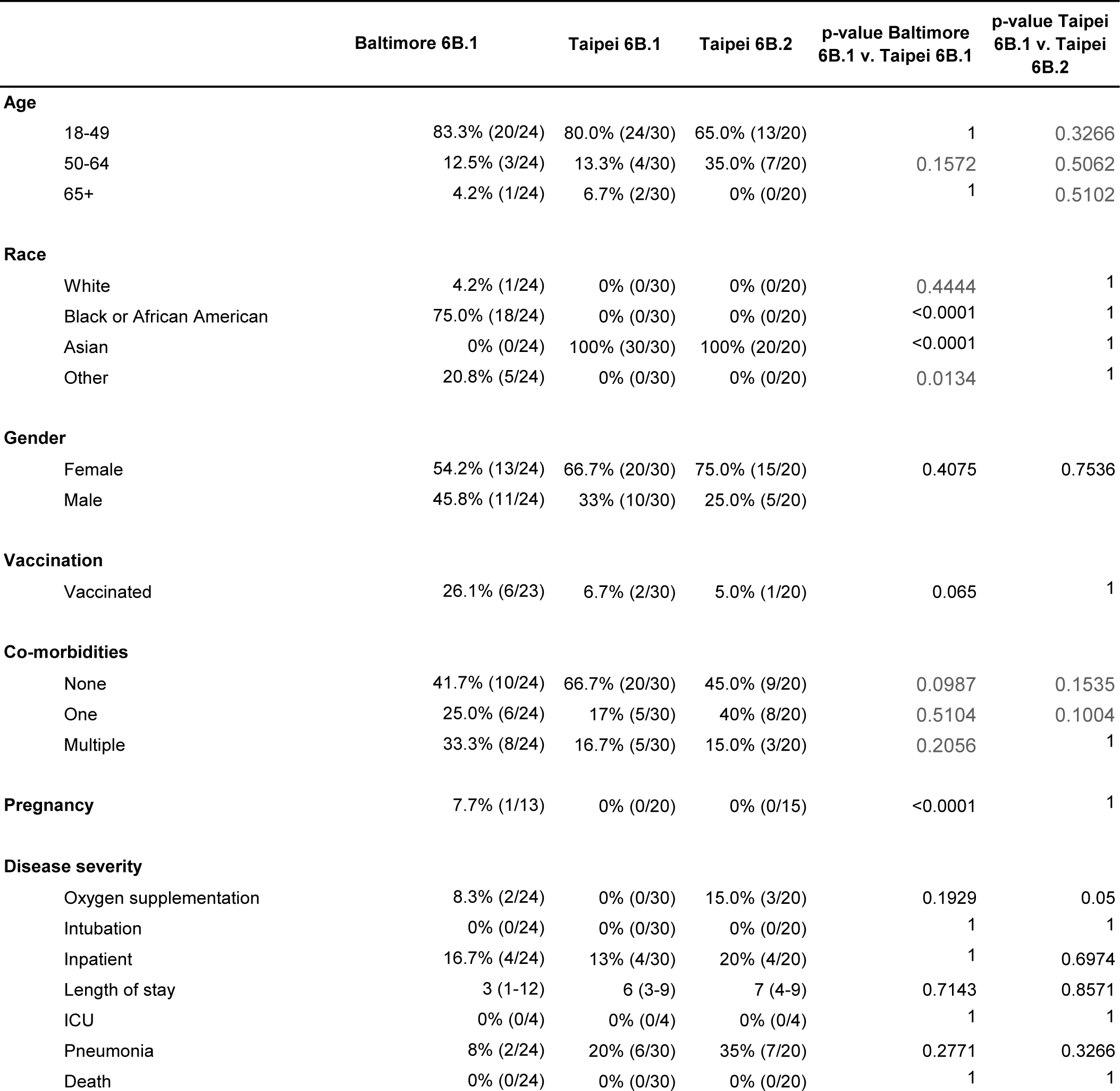
Clinical and demographic characteristics of H1N1 IAV-infected patients at Baltimore and Taipei based on HA clade

